# Community first response for cardiac arrest: comparing phased dispatch policies through Monte Carlo simulation

**DOI:** 10.1101/2024.01.17.24301457

**Authors:** Pieter L. van den Berg, Shane G. Henderson, Hemeng Li, Bridget Dicker, Caroline J. Jagtenberg

## Abstract

**Background:** Advanced Community First Responder (CFR) systems send so-called phased alerts: notifications with built-in time delays. The policy that defines these delays affects response times, CFR workload and the number of redundant CFR arrivals.

**Methods:** We compare policies by Monte Carlo Simulation, estimating the three metrics above. We bootstrap acceptance probabilities and response delays from 29,307 rows of historical data covering all GoodSAM alerts in New Zealand between 1-12-2017 and 30-11-2020. We simulate distances between the patient and CFRs by assuming that CFRs are located uniformly at random in a 1 km circle around the patient, for different CFR densities. Our simulated CFRs travel with a distance-dependent speed that was estimated by linear regression on observed speeds among those responders in the abovementioned data set that eventually reached the patient.

**Results:** The alerting policy has a large impact on the expected number of alerts sent, the redundant arrivals and the probability of patient survival. CFR app managers can use our results to identify a policy that displays a desirable trade-off between these performance measures.

## Introduction

Survival for out-of-hospital cardiac arrest (OHCA) can be significantly improved through bystander efforts.^1^ To shorten the time to good-quality cardiopulmonary resuscitation (CPR), some emergency call centers use mobile phone technology to rapidly locate and alert nearby trained volunteers. A number of such community first responder (CFR) systems are active in, for example, the United States,^2^ the United Kingdom^3^ and Europe.^4^

While details may differ between CFR systems, most systems alert multiple volunteers for each patient, and volunteers take some random amount of time to notice their alert and determine whether they will go. Volunteers subsequently indicate on the smartphone app whether they accept or reject the alert, and in case of acceptance, proceed to the patient’s location. Upon arrival, volunteers provide CPR until an ambulance takes over.

Since The International Liaison Committee on Resuscitation (ILCOR) identified a lack of literature on the effect of CFR systems in 2015,^5^ several papers have been published that investigate systems through surveys ^6, 2^ or retrospective data analysis^7^ (see Table 2 in that paper). Many reported a positive impact, e.g., statistically significant improved survival to hospital discharge^8^ or a lower degree of disability or dependence after survival.^9^ One study applied Monte Carlo simulation to compare four hypothetical CFR systems by estimating response times and patient survival.^10^

Challenges for an effective CFR system include the activation radius and volunteer density.^2^ One article studies densities of activated responders within a 1km circle and concludes that 10 volunteers /km^2^ are needed.^11^ A positive correlation between the fraction of inhabitants registered as volunteers and patient survival has been observed.^12^ It is reasonable to assume that managerial decisions for activation radii are dependent on volunteer density.

Despite the recent growth in literature on CFR systems, there is no study systematically evaluating different *alerting policies*. An alerting policy prescribes which volunteer to alert when. Simple alerting policies alert up to a fixed number of volunteers, and/or up to a fixed radius around the patient, and send all alerts at once. More complex alerting policies may use *phased* alerts, sending alerts in batches with time lags in between, to see if previous ones have been accepted. Such a policy is used by St John New Zealand, an ambulance provider that deploys the CFR system GoodSAM. St John New Zealand configured the system to dispatch CFRs in batches of 3 with time lags of 30 seconds.

For a single patient, it is best to send many alerts at once, but this leads to a higher volunteer workload as well as an increased likelihood of multiple volunteers arriving on scene, which may diminish their perceived contribution. Both aspects can lead to a long-term negative impact on volunteers’ willingness to respond. The question arises how the choice of alerting policy affects patient survival as well as volunteer fatigue.

We used Monte Carlo simulation to shed light on the question above, ultimately seeking to assist CFR system managers in understanding the consequences of their chosen alerting policy. We use historical data from GoodSAM responses in New Zealand between 2017 and 2020 to estimate travel times, as well as the probability of, and delay until, volunteer acceptance.

### 1.1. Preliminaries

The process of both CFR and EMS response is depicted in Figure 1. There, the name ‘EMS response time’ reflects the duration that is common for EMS providers to measure, and for which they often face targets and reporting obligations. For clarity, we have also added a ‘GoodSAM response time’, which starts at the moment the CFR system is activated.

**Figure 1:**
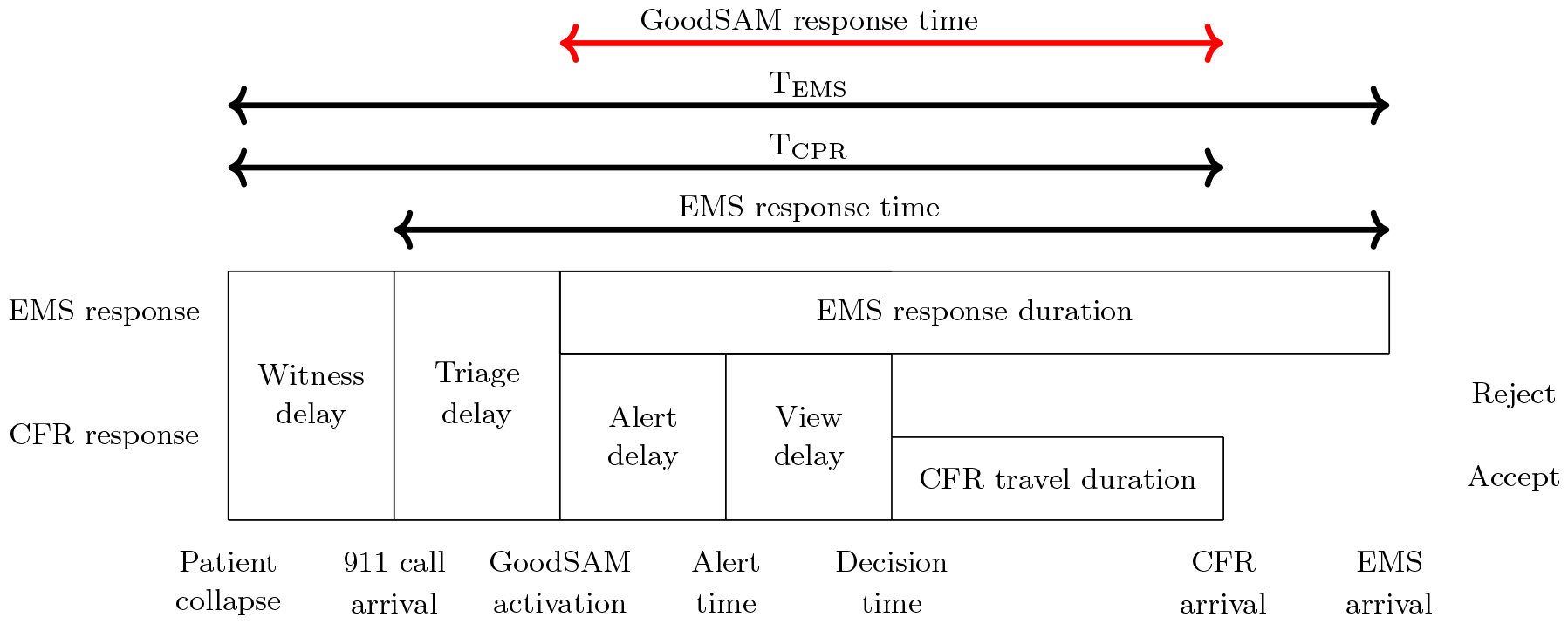
CFR response process.

EMS response times are measured from the moment of call arrival, so they are not ideal for estimating OHCA survival: the time that has passed since patient collapse is more relevant. To that end, we define another duration, T_EMS_: the duration between patient collapse and EMS arrival. Similarly, we define T_CPR_: the duration between patient collapse and the arrival of the first responder, regardless of whether this is a volunteer or EMS. This is summarized in Figure 1.

## 2. Material and methods

We applied Monte Carlo simulation to estimate the GoodSAM response time (see Figure 1) for selected dispatch policies. This involved bootstrapping delay and acceptance probabilities from historical records of GoodSAM responses in New Zealand, see Section 2.1. We report the estimated expected values of four key performance indicators (KPIs) described in Section 2.4.

### 2.1. Data

We used data obtained from the St John Ambulance Service, which operates the GoodSAM app in New Zealand. The data covers all OHCAs in the country between 1-12-2017 and 30-11-2020, along with their location and 911 call arrival time. The data also contains which CFRs were alerted through GoodSAM, at what time, and what their location was at the time of notification (*alert time* in Figure 1). Moreover, the data contains the time at which the volunteer reacted (*decision time*) by either accepting or rejecting the alert. Finally, the time volunteers arrived on scene (*CFR arrival*) is determined by the system, based on GPS signal. The data contains a total of 29,307 CFR alerts, which were either accepted (4009), accepted and dropped later (1199), rejected (7925), or not seen (16,174). From the 4009 accepting volunteers, 1776 eventually reached the patient.

For the view delay, we measured the duration between the alert time and the decision time. We ignored entries for which the alert time was equal to ‘nan’ (thereby excluding 1 entry). If the decision time was not listed, which is the case for not-seen alerts, we set the view delay to infinity. We treated both accepted and accepted-but-dropped-later entries as accepts, and we treated rejected and not-seen as rejects.This led to an empirical acceptance rate of 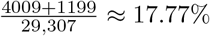. Figure 2 shows the empirical view delay distribution, as well as the probability of an accept, given a certain view delay. Not-seen alerts are not visible in this plot as these have an infinite view delay.

**Figure 2:**
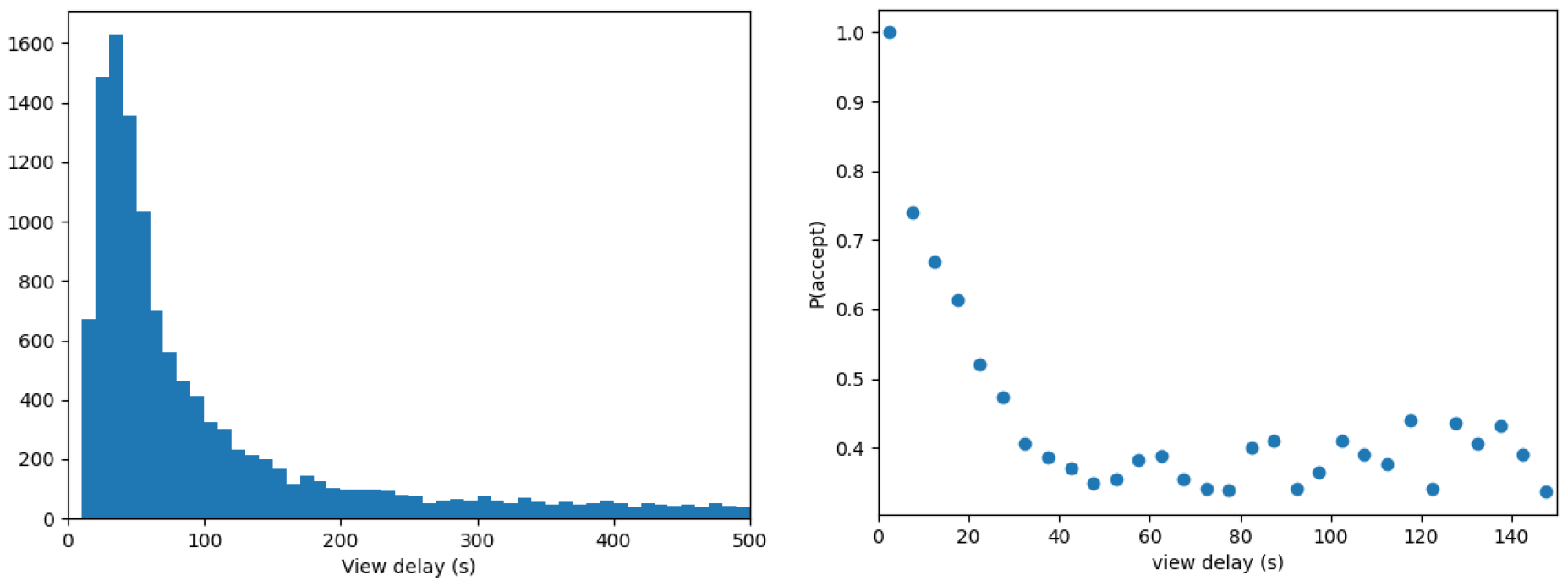
Historical view delays and acceptance probabilities as observed on GoodSAM in New Zealand.

Acceptance rates in Australia and New Zealand were found to be similar to those reported for other CFR smartphone apps.^6^

The CFR’s travel duration depends on their distance from the patient. Locations and travel times were only recorded for *alerted* CFRs, so we cannot observe information on non-alerted individuals. However, in our counterfactual simulations those CFRs *may* be alerted, so we estimate their distances using Monte Carlo simulation. For each CFR that is within 1km of the patient, we simulate their location uniformly at random in this circle.

To translate distance into travel time, we used empirical data on volunteer’s travel speed, conditioning on the distance between volunteer and patient. We excluded observations with a travel time of 0 seconds. We discretized the distances in 100-meter blocks and for each block, took the median speed of all volunteer responses in the data. We excluded the data for responses between 0 and 100 meters as we considered these less reliable. We then performed linear regression on the remaining medians to estimate the relationship between distance and speed.

### 2.2. Policies

We consider so-called alerting policies, which may base the decision to send an alert on the amount of time that has passed, and/or the responses that are received from previously alerted volunteers. Our policies only alert a more distant volunteer when all closer volunteers are already alerted. This helps to narrow down the set of all policies to realistic and promising ones. We further assume that no alerts are sent out after a volunteer accepts an alert, or 10 minutes have passed after GoodSAM activation, whichever comes first.

We evaluate the following policies:

1. *Send all at time 0* : alert all available volunteers within the dispatch radius immediately upon GoodSAM activation, which we call time 0.
2. *Send n at time 0* : Alert *n* volunteers immediately at time 0, and send no alerts after.
3. *Keep-n-active*: Alert *n* volunteers at time 0 and replace every incoming reject with an additional alert.
4. *NZ current policy* : Alert 3 volunteers at time 0 and 3 additional volunteers every 30 seconds until one volunteer accepts the alert, or 10 minutes have passed.

For *send n at time 0*, we consider the values of *n* between 1 and 15. For *keep-n-active* we consider values of *n* between 1 and 10. This gives a total of 27 policies.

### 2.3. Simulation

To estimate performance for the different policies, we applied Monte Carlo simulation, considering two sources of uncertainty: 1) the distance between the volunteers and the patient, and 2) the view delay and reply of alerted volunteers. The simulation evaluates 1,000 different OHCAs. For each OHCA, their CFR view delays and corresponding replies were bootstrapped 10,000 times from the empirical data. Each view delay and reply were jointly sampled to maintain the statistical dependence between the time a volunteer takes to respond and the likelihood that the reply is ‘accept’ (see Figure 2). This calculation estimates all KPIs with very narrow confidence intervals.

### 2.4. Performance metrics

We consider two KPIs related to the quality of care and two KPIs related to the inconvenience caused to volunteers.

#### Coverage

fraction of incidents that receive a CFR response within a given time threshold. We define this on the ‘GoodSAM response time’ (see Figure 1) and use a threshold of 5 minutes.

#### Lives saved

the number of OHCA patients that are expected to survive until hospital discharge, in New Zealand, annually. This number is estimated using the relationship between time to CPR, time to EMS (T_CPR_ and T_EMS_ in Figure 1) and the probability of survival as modeled by Waalewijn et al.^13^ We assumed a T_EMS_ of 13 minutes as this is St John New Zealand’s target for urgent requests.^14^ To estimate T_CPR_, we further need to estimate the ‘Witness delay’, which we assumed to be 1 minute, and the ‘Triage delay’, for which we used 124 sec (the median of the data).

The resulting survival percentage was converted to lives saved, by multiplying it by 5,141, which is the nationally observed number of OHCAs per year.^15^

#### Number of alerts

the average number of alerts sent per incident.

#### Redundant arrivals

the average number of volunteers arriving on scene after the first responder, per incident.

The relative performance of the different policies depends on the number of volunteers. This number depends on the density of volunteers in an area. We evaluated the different policies under a low (10 volunteers within 1km), medium (30 volunteers within 1km) and high (100 volunteers within 1km) density of volunteers. These densities roughly correspond to what would be found in Auckland, New Zealand if 0.2% of the inhabitants signed up (low), what is recommended in^11^ (medium) and our estimate for central Amsterdam (high).

## 3. Results

### 3.1 Parameter estimation

Figure 3a shows the observed median travel speeds for different distances, which increase with distance. Linear regression of travel speeds in km/h, *y*, on distances in meters, *x*, based only on the distances above 100 meters has an *R*^2^ of 0.96 and yields the relationship

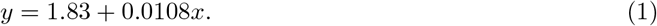

The resulting relationship between the distance in meters and travel time in minutes, plotted in Figure 3b, is

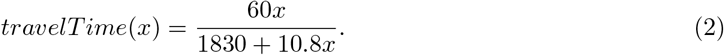

**Figure 3:**
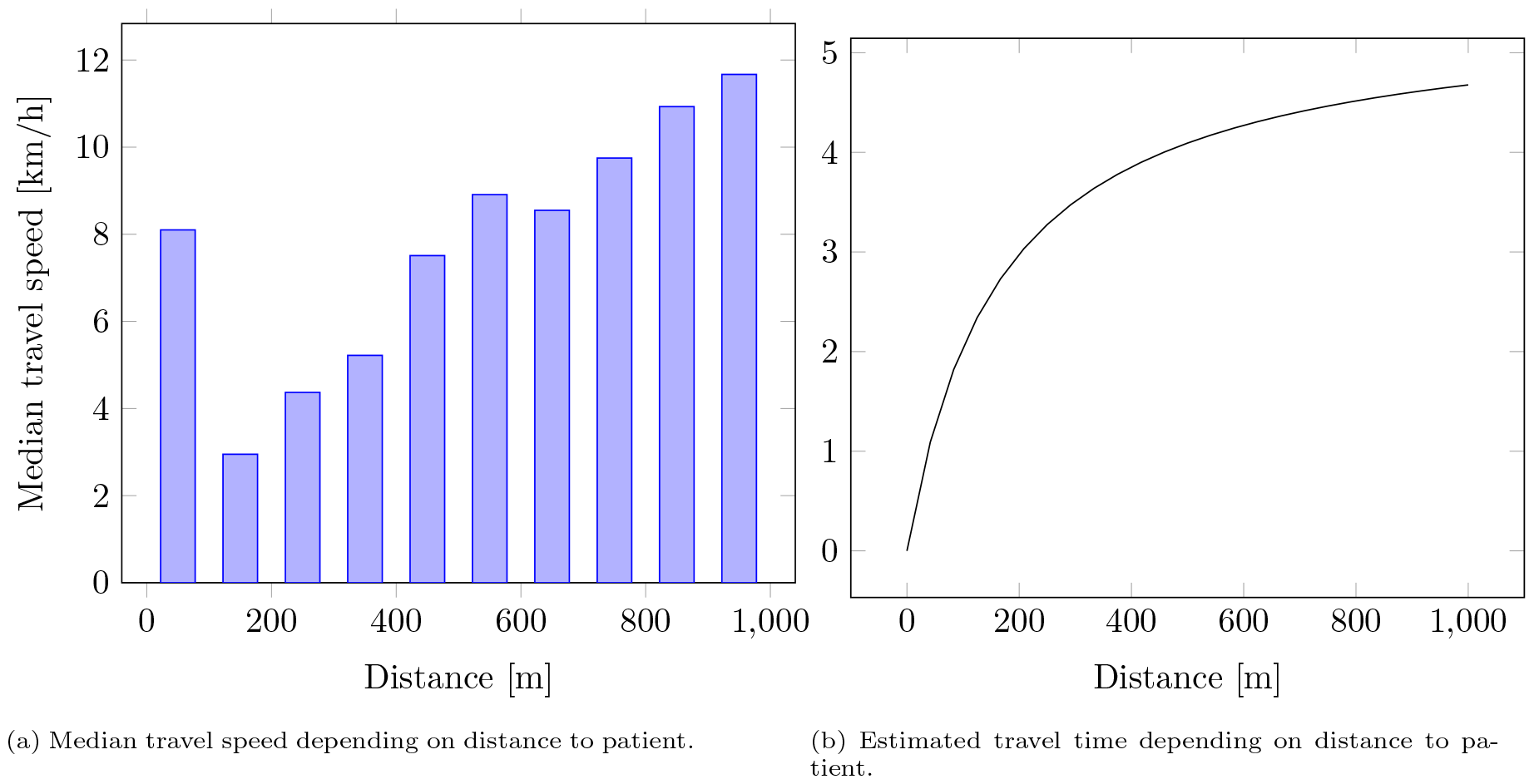
Distance-dependent travel times estimated based on historical data.

### 3.2. Performance of policies

Tables 1-3 show the results for the different policies for a low, medium and high number of volunteers. For reference, alerting 0 volunteers would lead to 98 survivors, 0 redundant arrivals, 0 alerts and 0 coverage.

**Table 1:**
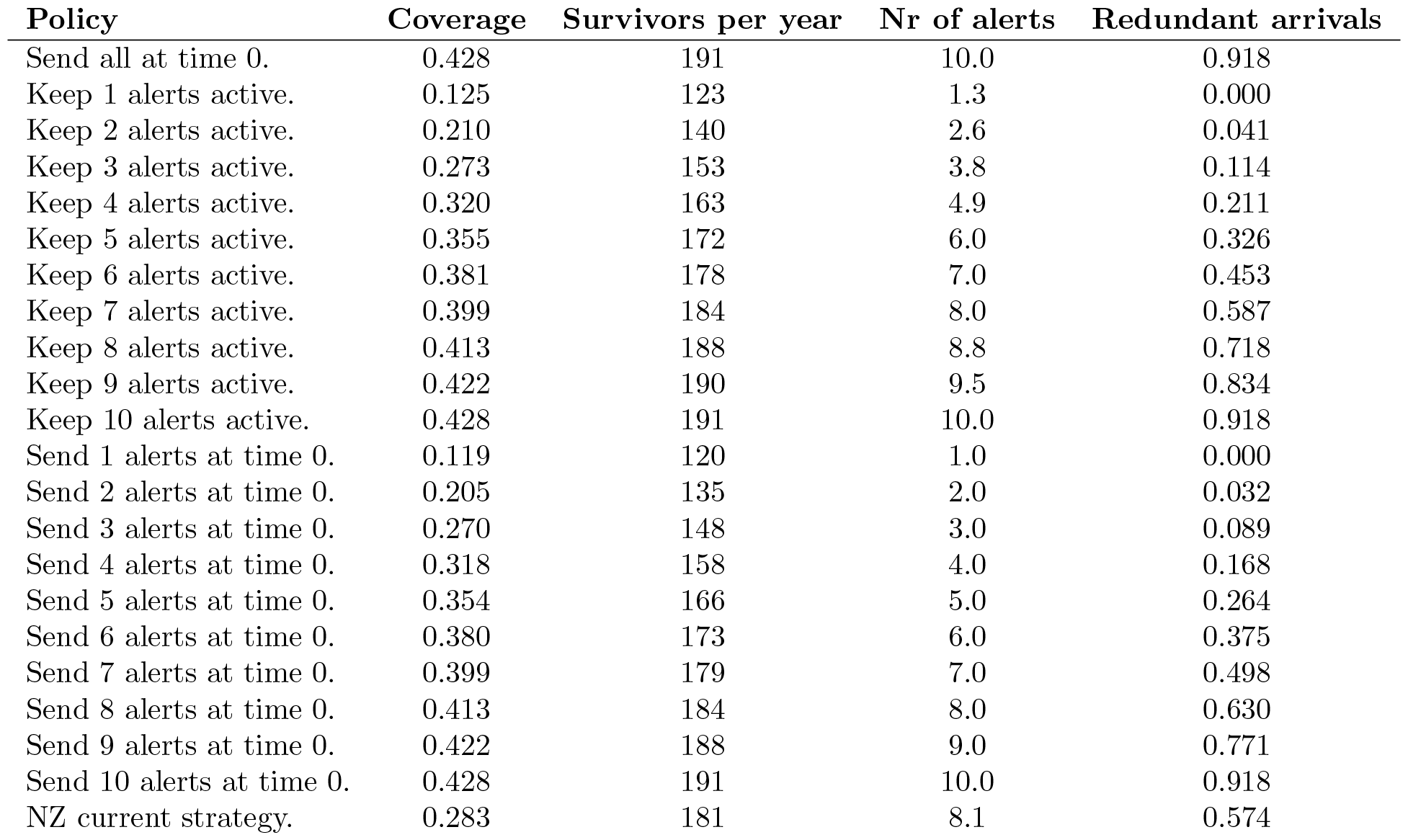
Results for *n* = 10 volunteers within 1 km, based on 1000 simulated volunteer locations, with 10000 view delay and acceptance simulations each. The maximum confidence interval halfwidth as a fraction of the mean among all these numbers was 0.013 (ignoring those cases where the mean was zero). Policies that send more than 10 alerts at time 0 are omitted, as there are only 10 eligible volunteers in this situation.

**Table 2:** Results for *n* = 30 volunteers within 1 km, based on 1000 simulated volunteer locations, with 10000 view delay and acceptance simulations each. The maximum confidence interval halfwidth as a fraction of the mean among all these numbers was 0.009 (ignoring those cases where the mean was zero.)

| Policy | Coverage | Survivors per year | Nr of alerts | Redundant arrivals |
| --- | --- | --- | --- | --- |
| Send all at time 0. | 0.806 | 231 | 30.0 | 4.333 |
| Keep 1 alerts active. | 0.149 | 128 | 1.3 | 0.000 |
| Keep 2 alerts active. | 0.258 | 148 | 2.6 | 0.041 |
| Keep 3 alerts active. | 0.343 | 164 | 3.8 | 0.114 |
| Keep 4 alerts active. | 0.411 | 176 | 4.9 | 0.211 |
| Keep 5 alerts active. | 0.468 | 185 | 6.0 | 0.326 |
| Keep 6 alerts active. | 0.516 | 193 | 7.1 | 0.455 |
| Keep 7 alerts active. | 0.556 | 199 | 8.1 | 0.595 |
| Keep 8 alerts active. | 0.591 | 205 | 9.2 | 0.743 |
| Keep 9 alerts active. | 0.620 | 209 | 10.2 | 0.897 |
| Keep 10 alerts active. | 0.646 | 213 | 11.2 | 1.056 |
| Send 1 alerts at time 0. | 0.134 | 124 | 1.0 | 0.000 |
| Send 2 alerts at time 0. | 0.240 | 142 | 2.0 | 0.032 |
| Send 3 alerts at time 0. | 0.326 | 157 | 3.0 | 0.089 |
| Send 4 alerts at time 0. | 0.398 | 169 | 4.0 | 0.168 |
| Send 5 alerts at time 0. | 0.457 | 178 | 5.0 | 0.264 |
| Send 6 alerts at time 0. | 0.507 | 186 | 6.0 | 0.375 |
| Send 7 alerts at time 0. | 0.550 | 193 | 7.0 | 0.498 |
| Send 8 alerts at time 0. | 0.586 | 199 | 8.0 | 0.631 |
| Send 9 alerts at time 0. | 0.617 | 203 | 9.0 | 0.771 |
| Send 10 alerts at time 0. | 0.643 | 207 | 10.0 | 0.918 |
| Send 11 alerts at time 0. | 0.666 | 211 | 11.0 | 1.071 |
| Send 12 alerts at time 0. | 0.686 | 214 | 12.0 | 1.228 |
| Send 13 alerts at time 0. | 0.703 | 216 | 13.0 | 1.389 |
| Send 14 alerts at time 0. | 0.718 | 218 | 14.0 | 1.552 |
| Send 15 alerts at time 0. | 0.731 | 220 | 15.0 | 1.719 |
| NZ current strategy. | 0.434 | 205 | 11.3 | 1.002 |

**Table 3:** Results for *n* = 100 volunteers within 1 km, based on 1000 simulated volunteer locations, with 10000 view delay and acceptance simulations each. The maximum confidence interval halfwidth as a fraction of the mean among all these numbers was 0.005 (ignoring those cases where the mean was zero.)

| Policy | Coverage | Survivors per year | Nr of alerts | Redundant arrivals |
| --- | --- | --- | --- | --- |
| Send all at time 0. | 0.996 | 259 | 100.0 | 16.768 |
| Keep 1 alerts active. | 0.168 | 134 | 1.3 | 0.000 |
| Keep 2 alerts active. | 0.297 | 159 | 2.6 | 0.042 |
| Keep 3 alerts active. | 0.400 | 178 | 3.8 | 0.114 |
| Keep 4 alerts active. | 0.484 | 193 | 4.9 | 0.211 |
| Keep 5 alerts active. | 0.552 | 205 | 6.0 | 0.326 |
| Keep 6 alerts active. | 0.609 | 214 | 7.1 | 0.456 |
| Keep 7 alerts active. | 0.657 | 222 | 8.1 | 0.595 |
| Keep 8 alerts active. | 0.698 | 228 | 9.2 | 0.743 |
| Keep 9 alerts active. | 0.733 | 233 | 10.2 | 0.897 |
| Keep 10 alerts active. | 0.763 | 237 | 11.2 | 1.056 |
| Send 1 alerts at time 0. | 0.145 | 129 | 1.0 | 0.000 |
| Send 2 alerts at time 0. | 0.263 | 152 | 2.0 | 0.032 |
| Send 3 alerts at time 0. | 0.362 | 170 | 3.0 | 0.089 |
| Send 4 alerts at time 0. | 0.445 | 184 | 4.0 | 0.168 |
| Send 5 alerts at time 0. | 0.515 | 196 | 5.0 | 0.265 |
| Send 6 alerts at time 0. | 0.575 | 205 | 6.0 | 0.375 |
| Send 7 alerts at time 0. | 0.626 | 214 | 7.0 | 0.498 |
| Send 8 alerts at time 0. | 0.670 | 220 | 8.0 | 0.631 |
| Send 9 alerts at time 0. | 0.708 | 226 | 9.0 | 0.771 |
| Send 10 alerts at time 0. | 0.740 | 231 | 10.0 | 0.918 |
| Send 11 alerts at time 0. | 0.769 | 235 | 11.0 | 1.071 |
| Send 12 alerts at time 0. | 0.794 | 238 | 12.0 | 1.228 |
| Send 13 alerts at time 0. | 0.815 | 241 | 13.0 | 1.388 |
| Send 14 alerts at time 0. | 0.834 | 244 | 14.0 | 1.552 |
| Send 15 alerts at time 0. | 0.851 | 246 | 15.0 | 1.718 |
| NZ current strategy. | 0.615 | 229 | 11.4 | 1.021 |

By only alerting the closest volunteer, the number of survivors can be increased to 120, 124, or 129, for 10, 30 or 100 volunteers within 1km, respectively. For coverage, the contribution of the closest volunteer is 11.9%, 13.4%, or 14.5%, respectively.

Comparing policies *Send n alerts at time 0*, with *Keep n alerts active* indicates that a similar level of survival can be obtained with 1 alert less at time 0, by replacing rejects with additional alerts. This would reduce the impact of the alerts on the volunteers (both in terms of redundant arrivals as well as the number of alerts) without sacrificing survival.

The current policy used by GoodSAM in New Zealand on average leads to sending out 11.4 alerts per incident, if sufficiently many volunteers are within 1km from the patient. The corresponding number of expected survivors per year is 181, 205, or 229 for a low, medium and high number of volunteers. Coverage as a result of this policy is estimated at 28.1%, 43.6%, or 61.5%, respectively.

The policy that sends an alert to all volunteers at time 0 by definition yields the highest estimated survival (191, 231 and 259 survivors per year) and coverage (42.3%, 80.7% and 99.6%), but also gives the highest estimated number of redundant arrivals (0.919, 4.333 and 16.77) and alerts.

## 4. Discussion

The trade-offs between the KPIs in Tables 1-3 help a CFR system manager to identify a desirable dispatch policy. This choice will likely depend on the volunteer density in the region. Our reported number of volunteers within 1 km of the patient has a direct relationship with the volunteer density. The three cases we considered correspond to a density of 3.18 vol/km^2^, 9.55 vol/km^2^, and 31.83 vol/km^2^, respectively. For reference, the literature recommends a volunteer density of at least 10 vol/km^2^.^11^

The insight that a different density might lead to a different preferred policy might be somewhat new in the academic literature; however, it was already known to GoodSAM. For this reason, their system offers the ability to configure different rules for urban and rural areas, which is done by uploading KML files that represent parts of the map. This feature implies that potential insights from this paper can readily be implemented in practice.

Alerting the same number of volunteers leads to higher expected survival and better coverage as volunteer density increases. This effect is visible throughout Tables 1-3, and also holds in generality, assuming all other system parameters remain equal. The reason is that the *n*-th closest volunteer is expected to be nearer to the patient when volunteer density is higher.

Comparing policies *Send 6 alerts at time 0* and *Send 10 alerts at time 0* shows different survival numbers for different volunteer densities. In Table 1 the difference is 13 lives, while in Table 3 it is 24 lives. The reason is that in Table 1, the 6th-10th volunteers are farther from the patient than they are in Table 3.

Perhaps more surprising is that the policy *Send 6 alerts at time 0* gives approximately the same survival probability as *Keep 5 alerts active*, and that this is the case in each of the three tables. We have no intuition for this, but conjecture that this result would be found for any volunteer density within the entire range of 3.18 – 31.83 vol/km^2^.

The current strategy in New Zealand of alerting volunteers in batches of three with 30-second intervals leads to few redundant arrivals, but at a low volunteer density, this comes at a high cost in terms of coverage. This insight might be a reason to fine-tune the number of alerts per batch and the interval between batches to better align with the view delay and acceptance probability, especially in sparsely populated parts of the country.

A limitation of this study is that the time on-scene is based on the geolocation of the phone which may have some error in it due to gaps in cell tower coverage and the resulting inability to ping the phone when it is in the vicinity of the patient. It is unknown how often this situation occurs. The stop-gap to this is that responders can also manually generate a time on scene by pressing a button on the app, but responders do not always do this.

Another limitation is that we focused on volunteers who directly go to the patient to provide CPR. However, in practice, certain CFR systems differentiate alerts, directing some volunteers to immediately attend to the patient and others to retrieve an Automated External Defibrillator (AED). Such a system raises new questions; answering those questions would require an extension of our methods.

We limited attention to policies that alert far-away volunteers only if all nearer ones have already been alerted. While this choice may seem restrictive, these policies are optimal when we do not have volunteer-specific knowledge. While some CFR systems collect volunteer-specific parameters such as acceptance rates, to the best of our knowledge, no system currently uses this information in their real-time dispatch decisions.

We explored results for three different volunteer densities, but a CFR system will have varying volunteer densities throughout the day and the region. It might then be worthwhile to differentiate the dispatch policy within one system through times of the day and parts of the region. Accurately evaluating such a CFR system would require running simulations with the correct mix of densities and policies, thereby introducing complexity and likely diminishing insight. Therefore we decided not to pursue that approach here.

Although it is plausible that a high number of alerts and/or redundant arrivals reduce future CFR engagement, there is, to the best of our knowledge, no study that quantifies the magnitude of this effect. We welcome such studies, as they might aid in designing even better dispatch policies. The current knowledge gap led us to refrain from recommending a specific dispatch policy, instead presenting a list of options from which a CFR system manager can choose.

Future research might address more sophisticated policies. For example, we conjecture that it would be a good idea to consider a CFR as inactive when they have not responded for some prescribed time. Moreover, it could be useful to let the chosen policy depend on the real-time locations of volunteers and the time of day. If, for example, the third volunteer is far from the patient, it might be better to alert only two, and more alerts might be sent at times when volunteers are less likely to respond.

## 5. Conclusions

A CFR system’s dispatch policy affects the trade-off between the number of alerts, redundant arrivals, and patient survival. In comparison to sending all alerts immediately, it is beneficial to send a reduced number of alerts immediately plus additional alerts upon receiving rejections. This reduces the number of redundant arrivals as well as the number of alerts, without losing much in terms of patient survival. The benefit of such a policy increases with an increasing number of volunteers around the patient.

This paper quantified KPI trade-offs for several reasonable dispatch policies and varying volunteer densities, and hence can guide CFR system managers in choosing a policy that suits their region. The choice likely depends on the volunteer density in the region and might even be differentiated within one system if volunteer densities vary over the day and between different parts of the region.

We hope that this article will stimulate discussion on phased dispatching of CFRs and pave the way for incorporating methods from the field of operations research in designing improved dispatch policies.

## Data Availability

Data subject to third party restrictions. Data available from the authors with the permission of St John New Zealand

## Acknowledgements

We thank GoodSAM and St John Ambulance Service New Zealand for access to data.

